# Multi-omics data integration to identify metabolism pathways and therapeutic targets for cardiac disease

**DOI:** 10.1101/2024.02.27.24303421

**Authors:** Sophie C. de Ruiter, Marion van Vugt, Chris Finan, Rui Providencia, Sandesh Chopade, Diederick E. Grobbee, Hester M. den Ruijter, Sanne A.E. Peters, A. Floriaan Schmidt

**Affiliations:** Julius Center for Health Sciences and Primary Care, University Medical Center Utrecht, Utrecht University, Utrecht, the Netherlands; Department of Cardiology, Amsterdam Cardiovascular Sciences, Amsterdam University Medical Centres, University of Amsterdam, Amsterdam, the Netherlands; Institute of Cardiovascular Science, Faculty of Population Health, University College London, London, United Kingdom; Division Heart and Lungs, Department of Cardiology, University Medical Center Utrecht, Utrecht University, Utrecht, the Netherlands; UCL British Heart Foundation Research Accelerator, London, United Kingdom; Health Data Research UK and Institute of Health Informatics, University College London, London, United Kingdom; Barts Heart Centre, St Bartholomew’s Hospital, Barts Health NHS Trust, London, United Kingdom; Laboratory of Experimental Cardiology, Division Heart and Lungs, University Medical Center Utrecht, Utrecht University, Utrecht, the Netherlands; The George Institute for Global Health, School of Public Health, Imperial College London, London, UK

## Abstract

**Introduction:** Urinary breakdown products, representing kidney regulated filtration of metabolism end products, contain cardiac disease biomarkers such as NT-proBNP. We set out to integrate plasma proteins with metabolism pathways, as reflected by urinary breakdown products, to identify potentially druggable metabolism pathways for cardiac disease.

**Methods:** Data was leveraged from a genome-wide association study (GWAS) on 954 urinary breakdown products. Mendelian randomisation was used to identify urinary breakdown products associating with atrial fibrillation (AF), heart failure (HF), dilated cardiomyopathy (DCM), or hypertrophic cardiomyopathy (HCM). By interrogating eight independent plasma protein GWAS, jointly including 92,277 participants and 1,562 unique proteins, we identified druggable plasma proteins with a directionally concordant effect on urinary breakdown products and cardiac outcomes.

**Results:** In total, 38 unique urinary breakdown products associated with cardiac disease, predominantly covering breakdown products from amino acid metabolism (n=12), xenobiotic metabolism (n=5), and unclassified metabolism origins (n=16). Subsequently, we identified 32 druggable proteins expressed in cardiac tissue, which had a directionally concordant association with the identified urinary breakdown products and cardiac outcomes. This included positive control findings, for example higher values of AT1B2 (targeted by digoxin) decreased the risk of HCM, which we were able to link to a novel unclassified urinary breakdown product (X-15497). Additionally, we showed that increased plasma RET values, a mediator of GDF-15 signalling, reduced the risk of HF, and linked this to the novel unclassified urinary breakdown product X-23776.

**Conclusion:** We have identified amino acid, xenobiotic and unclassified metabolism as important pathways contributing to cardiac disease and prioritised 32 druggable proteins as potential therapeutic targets.

## Introduction

Heart failure (HF) and atrial fibrillation (AF) are cardiac diseases which manifest as ventricular and atrial dysfunction, respectively. While AF frequently affects people without established HF, in people with HF an AF diagnosis often signals early progressive HF, where ventricular disfunction has an early knock-on effect on atrial rhythm and output. People with hypertrophic cardiomyopathies (HCM), a subtype of HF characterized by an abnormally large and stiff cardiac septum, which may limit cardiac output and affect diastolic function, are particularly susceptible to develop AF. Aside from HCM, dilated cardiomyopathy (DCM) represents a second major non-ischemic HF subtype, where the left ventricle becomes enlarged and thin, limiting cardiac contractility. As such, AF and HF are closely interrelated diseases, with specific underlying pathologies like HCM and DCM playing distinct roles in their manifestation.

Aside from N-terminal prohormone of brain natriuretic peptide (NT-proBNP), which associates with HF and AF progression(1–3), there is paucity of biomarkers to assist medical practitioners in tailored treatment management, diagnosis, and prognosis. NT-proBNP, which is secreted by cardiomyocytes (cardiac cells responsible for contraction), can be measured in both blood plasma and urine, illustrating the interface between cardiac tissue processes, blood, and urine. Urine biomarkers provide a non-invasive measurement of potential causal metabolic changes reflecting disease onset and progression. While urinary breakdown products represent by-products cleared from plasma, the source of these breakdown products may reflect pathophysiological changes in tissues such as the cardiac muscle. Cardiac function and kidney function are intertwined, through for example the renin-angiotensin-aldosterone system, and the concept of cardiorenal syndrome has been implemented to describe disorders involving both organs, with dysfunction in one organ causing acute or chronic dysfunction in the other(4, 5). Urinary breakdown products are therefore a potentially important source of non-invasive disease markers, which may unveil upstream involvement of biological pathways relevant for cardiac progression and/or novel cardiac therapeutics.

Recently, availability of affordable, high-throughput assays capable of measuring hundreds of proteins and metabolites has facilitated their analysis in large sample size genome wide association studies (GWAS). Given that proteins form the main constituents of drug targets, GWAS on protein values provide an invaluable resource to identify potential drug targets through genomics tools such as Mendelian randomisation (MR). Utilizing MR, one can infer the effect of an exposure (e.g. a breakdown product or protein) on the risk of disease, by identifying genetic variants strongly associated with the exposure, and determining whether there is a dose-response relationship between the genetic associations with the exposure and the genetic associations with disease(6, 7). Given that genetic variants are fixed at gametogenesis, genetic evidence is robust against bias due to confounding or reverse causation(8, 9). These principles have been extensively validated in the context of cardiovascular disease(10–13), including in a recent study on the effects of plasma proteins on cardiac function and structure(14).

Although urinary breakdown products are important markers of potential disease mechanisms, they are not directly actionable because they have already been excreted. Plasma proteins are however still actionable and are the main constituents of drug targets. Therefore, we integrated information from urinary metabolism breakdown products to identify metabolism pathways relevant for cardiac disease and information from plasma proteins to identify drug targets that may be used to intervene in these pathways.

In the current manuscript we used MR to analyse urinary metabolism breakdown products to identify metabolism pathways with possible involvement with four cardiac disease traits: AF, HF, DCM, and HCM. Subsequently, we sought to establish potential plasma protein intermediates by identifying proteins associating with urinary breakdown product values, as well as with cardiac disease risk. The Human Protein Atlas (HPA) was consulted to identify proteins expressed in cardiac tissue, providing further support for cardiac involvement. The subset of proteins expressed in cardiac tissue were subsequently mapped to biological pathways, with druggability determined through linkage with ChEMBL.

## Methods

### Available data

We used a GWAS on 954 urinary breakdown products, which were identified and quantified among 1,627 individuals using a non-targeted mass spectrometry assay provided by Metabolon(15). This untargeted approach allows for comprehensive detection of breakdown products, including those of unknown origin. Breakdown products were categorised based on the metabolic origin of the urinary breakdown products: amino acids, carbohydrates, cofactors and vitamins, energy, lipids, nucleotides, peptides, xenobiotics, and unclassified. Xenobiotic breakdown products - substances not naturally produced by the human body, include breakdown products from drugs, environmental pollutants, and various synthetic compounds. Unclassified breakdown products represent potentially novel biomarkers, as they have not yet been related to an established metabolic origin. These breakdown products were noted for their recurring chromatographic and spectral properties and have been assigned unique COMP-IDs and/or CHEM-IDs (**Supplementary Table S1**), which allows for identification and comparison in future studies and by other analysis platforms. Genetic associations with plasma protein values were available from the eight GWAS, detailed in Supplementary Note 1. Genetic associations with cardiac disease onset were available from Nielsen *et al*. (60,620 AF cases)(16), from Shah *et al*. (47,309 HF cases)(17), from Garnier *et al*. (2,719 DCM cases)(18), and from Zhou *et al.* (2,993 HCM cases)(19). Study specific participants characteristics and case/control definitions are described in Supplementary Note 1. All studies obtained informed consent from participants and complied with the Declaration of Helsinki.

### Mendelian randomisation analyses

Three MR analyses were conducted: 1) genome-wide MR identifying metabolism pathways which, as reflected by urinary breakdown products, associate with onset of cardiac disease (step 1 **Figure 1, Appendix Tables S1, S2**), 2) *cis*-MR identifying plasma proteins affecting values of urinary breakdown products (step 2 **Figure 1**), and 3) *cis*-MR identifying plasma proteins which associate with the onset of cardiac disease (step 3 **Figure 1**). In the genome-wide MR we selected genetic variants from across the genome, while in the *cis*-MR genetic variants were selected from a 200 kilobase pair window around and within the protein encoding gene. In both cases, variants were selected based on an exposure F-statistic of at least 24 and a minor allele frequency of 0.01 or larger. The remaining variants were clumped to a linkage disequilibrium (LD) r-squared of 0.30, based on a random 5,000 sample of unrelated UK Biobank participants. The *cis-*MR analyses (steps 2-3 **Figure 1)** were conducted per individual protein GWAS, where the largest sample size GWAS was used in the case of overlap in measured proteins.

**Figure 1.**
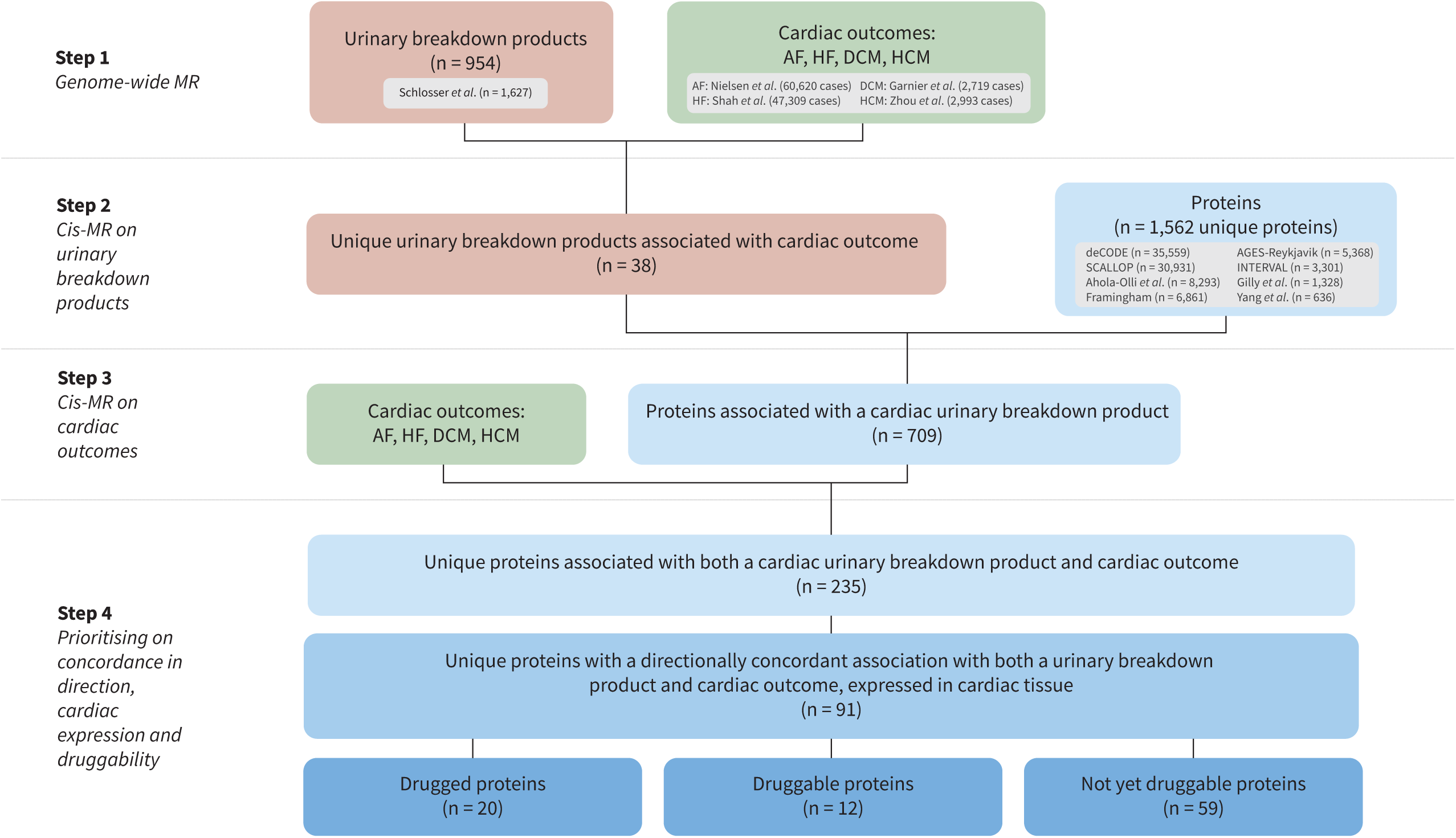
Flowchart of the main steps of this study. Abbreviations: AF = atrial fibrillation, DCM = dilated cardiomyopathy, HCM = hypertrophic cardiomyopathy, HF = heart failure.

MR analyses were conducted using generalized least squares (GLS) implementations of the inverse variance weighted (IVW) estimator, as well as the pleiotropy robust MR-Egger estimator(20). The GLS implementation was used to additionally correct for residual LD, which optimized estimator precision(21). To reduce the potential for horizontal pleiotropy, we excluded variants with large leverage statistics (larger than three times the mean leverage) and/or outlier statistics (Chi-square larger than 10.83). Analyses with fewer than 6 variants as instrument were discarded to ensure we had sufficient data to accurately model the exposure effects. The potential influence of horizontal pleiotropy was further minimized by applying a model selection framework identifying the MR model (IVW or MR-Egger) most supported by the available data (Supplementary Note 1).

### Effect estimates and multiple testing

MR results are presented as mean differences (MD) for continuous outcomes, or odds ratios (ORs) for binary outcomes, representing the effect of an SD increase of the exposure (i.e., either urinary breakdown product or plasma protein value). Effect estimates are not constrained to be positive (i.e., they can indicate an increase or decrease in the outcome) and are accompanied by 95% confidence intervals (CIs) and p-values. Breakdown product effects on disease were filtered for a multiplicity corrected p-value threshold of 1.37×10LL based on the 365 principal components that were needed to explain 90% of the urinary breakdown product variance (**Appendix Figure S1**). Protein MR effect estimates were evaluated against a multiplicity corrected p-value of 8.42×10^-7^ for the breakdown products, based on the number of tested proteins (n=1,562) and breakdown products (n=38). The multiplicity corrected p-value was 1.76×10^-5^ for cardiac outcomes, based on the number of tested proteins (n=709) and outcomes (n=4). Proteins are referred to using their Uniprot label(22), while genes are referred to using their Ensembl label in italicised font.

### Triangulation of evidence through consideration of concordant cardiac effects

The three distinct MR analyses were integrated by I) identifying urinary breakdown products affecting cardiac disease, II) identifying whether there were proteins associated with these urinary breakdown products, and III) focussing on the subset of proteins with a directionally concordant effect on the same cardiac disease trait the urinary breakdown products associated with. For example, if a higher value of a urinary breakdown product decreased the risk of HF, and the protein increased the urinary breakdown product value, the protein effect was considered directionally concordant if in turn higher protein values decreased HF risk. Conversely, if a higher value of a urinary breakdown product increased the risk of HF, and the protein decreased the urinary breakdown product value, the protein effect would still be considered directionally concordant if higher protein values decreased HF risk.

When all these conditions are met – directional concordance between the protein, breakdown product, and cardiac outcome – we describe this as a triangulated association (**Figure 2**). We note that because each performed MR analysis makes a distinct, and independent, assumption on the absence of horizontal pleiotropy, focusing on the directionally concordant results is anticipated to identify a robust subset of analyses with limited influence of potential horizontal pleiotropy.

**Figure 2.**
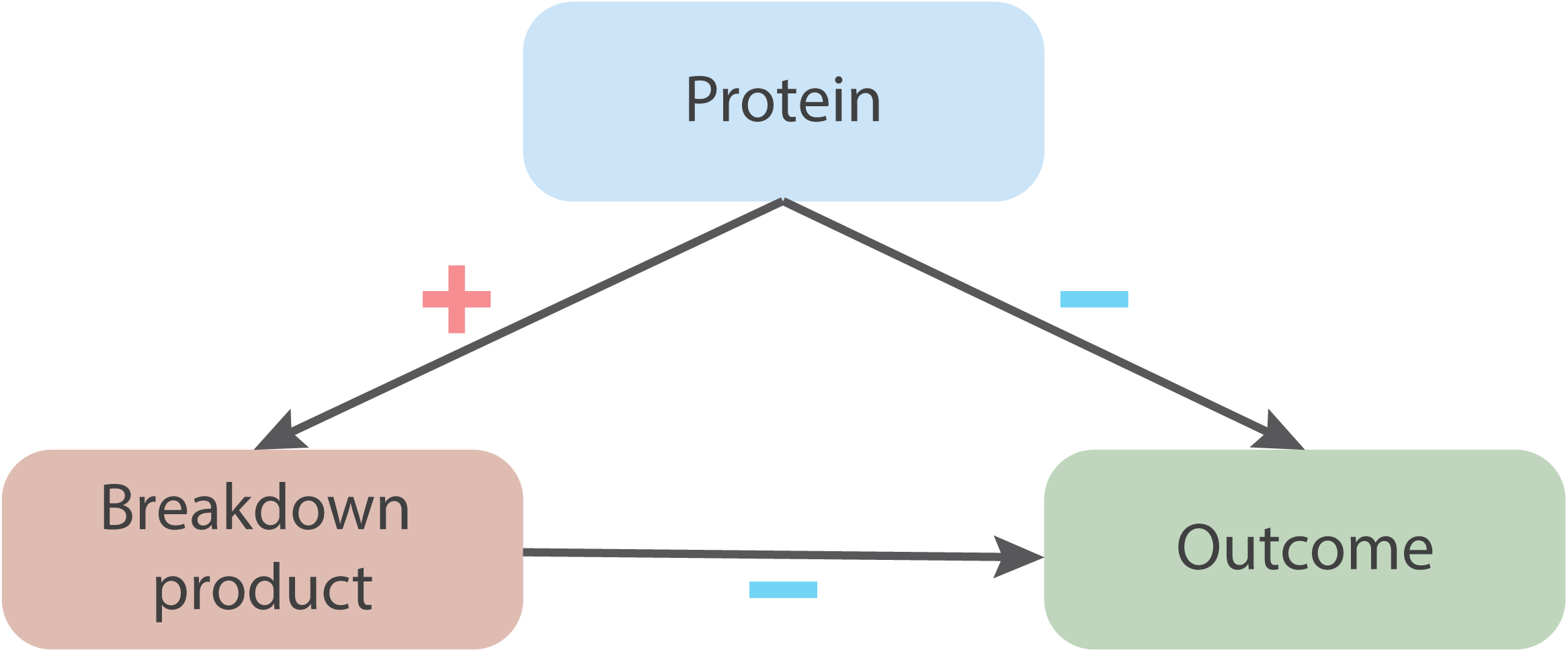
A triangulation diagram illustrating an example of a triangulated association – with directional concordance – between a protein and its effects on urinary breakdown product values and cardiac disease. N.B. The plus symbol (in red) represents a positive association. The minus symbol (in blue) represents a negative association (a risk-decreasing effect). Directional concordance is achieved when the direction of effects aligns consistently, whether increasing or decreasing.

### Prioritisation of proteins expressed in cardiac tissue

The set of proteins with a triangulated effect on breakdown product and cardiac outcome were subsequently prioritised for the presence of cardiac tissue mRNA expression, differentiating between proteins with general cardiac expression, and proteins which were overexpressed (Supplementary Note 1). We also tested for overexpression in other tissues, providing greater context regarding the type of protein.

### Annotation of prioritised proteins

We identified plasma proteins targeted by approved drugs (drugged proteins), as well as potentially druggable proteins based on ChEMBL-v33. For the drugged and druggable proteins, we additionally extracted compound indications and side-effects, sourcing information from ChEMBL and the British National Formulary (BNF, access date September 4, 2021). Finally, all proteins available in the considered GWAS were mapped to biological pathways using the Reactome knowledgebase-v85(23) and prioritised proteins (combined irrespective of cardiac or metabolite class association, as well as, stratified by cardiac disease, or stratified by metabolite class) were tested for enrichment against the full set of 1,567 proteins.

### Partial replication of protein associations with cardiac traits

Due to the availability of 8 proteomics GWAS, some studies assayed the same proteins, which we leveraged to replicate our findings, specifically focussing on associations with cardiac traits. Replication was sought using a nominally significant p-value of 0.05 or smaller and considering the effect direction of the triangulated analysis. In addition, a more stringent p-value threshold of 8.93×10^-4^ (0.05 divided by the number of proteins available in more than one study) was used for more conservative replication.

## Results

### Urinary breakdown products associating with the onset of cardiac disease

We conducted Mendelian randomisation to identify urinary breakdown products linked to cardiac disease, unveiling potential non-invasive biomarkers and altered metabolism pathways involved in cardiac disease progression. Out of 954 evaluated urinary breakdown products (**Appendix Table S1**), 38 breakdown products were found to be associated with at least one type of cardiac disease: 25 for AF, 11 for HF, 5 for DCM, and 14 for HCM (step 1 **Figure 1**, **Appendix Tables S2, S3, Figure 3**). Urinary breakdown products were frequently associated with AF, where particularly amino acid and xenobiotic breakdown products associated with cardiac diseases. For example, higher values of 1-methylhistamine increased the risk of HF (OR 1.07, 95%CI 1.05; 1.09), while decreasing the risk of AF (OR 0.83, 95%CI 0.76; 0.90). Higher values of 1-methylxanthine, a xenobiotic breakdown product of caffeine, decreased the risk of AF, DCM, and HCM (**Appendix Table S2, Figure 3)**. Finally, we were able to associate 16 urinary breakdown products of unknown metabolic origin (unclassified breakdown products) with cardiac disease.

**Figure 3.**
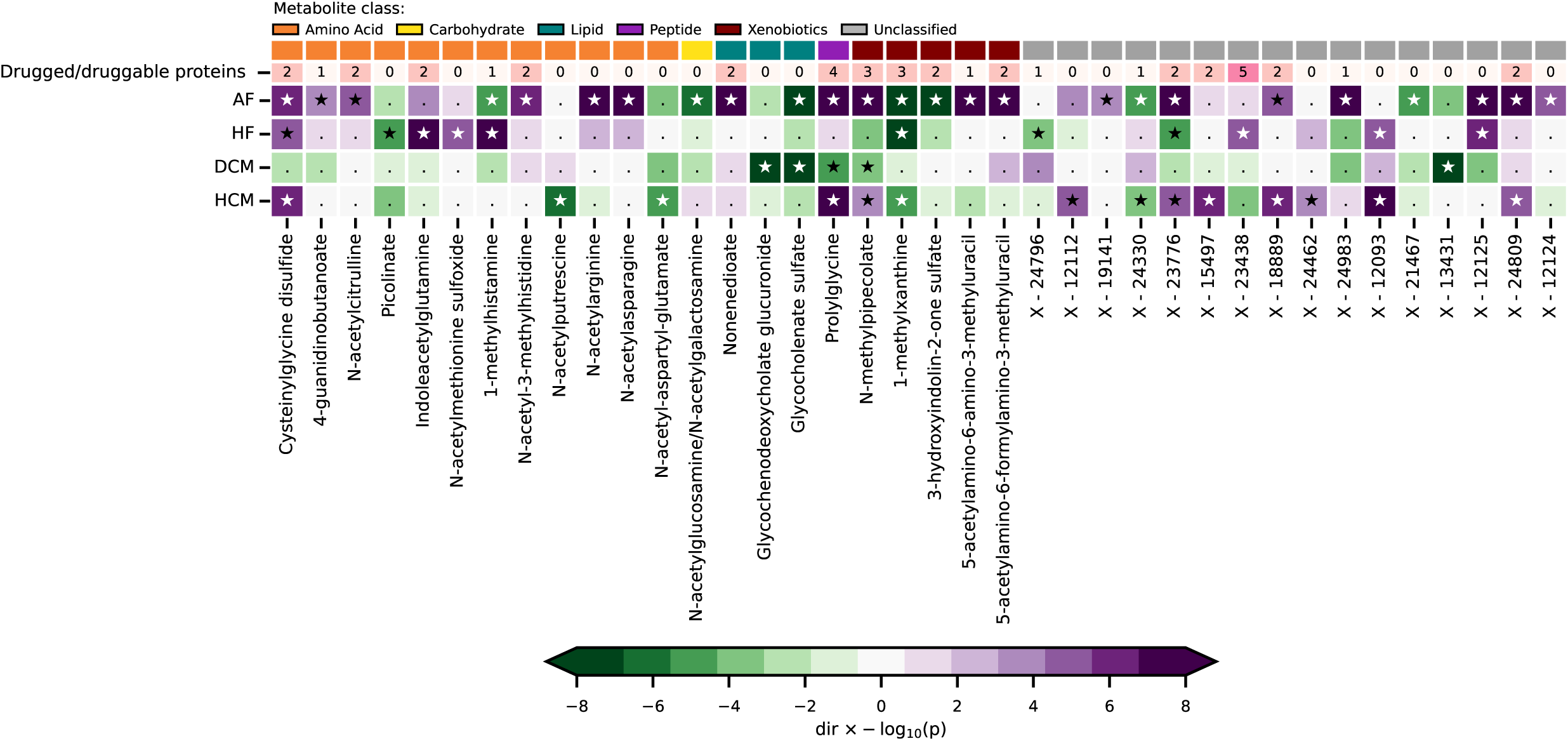
Urinary breakdown products associated with at least one cardiac outcome, presented per cardiac outcome. N.B. Presented p-values are corresponding to effect estimates obtained from Mendelian randomisation analyses leveraging genetic data on urinary breakdown products and cardiac diseases. Significant associations after multiplicity correction are indicated by a star, non-significant associations by a dot. The top row provides the number of drugged and druggable proteins, sourced from BNF and ChEMBL. Genetic associations with the cardiac outcomes were obtained from Nielsen *et al*. (60,620 AF cases)(16), from Shah *et al*. (47,309 HF cases)(17), from Garnier *et al*. (2,719 DCM cases)(18), and from Zhou *et al.* (2,993 HCM cases)(19). Genetic associations with urinary breakdown products are obtained from Schlosser et al.(n=1,627)(15). For more detailed information, please refer to the Methods section and **Appendix Table S2**. Abbreviations: AF = atrial fibrillation, DCM = dilated cardiomyopathy, HCM = hypertrophic cardiomyopathy, HF = heart failure, MR = Mendelian randomisation.

### Triangulated proteins with concordant effects on urinary breakdown products and cardiac diseases

Mendelian randomisation was used to determine associations between plasma proteins and the 38 urinary breakdown products associated with cardiac disease. We identified 709 proteins associated with at least one of the 38 urinary breakdown products (step 2 **Figure 1, Appendix Figure S2**). We then used MR to identify which of these proteins also associated with the considered cardiac diseases and identified 235 proteins (step 3 **Figure 1, Appendix Figure S3**). By considering directional concordance and cardiac tissue expression we identified 91 prioritised proteins (step 4 **Figure 1, Appendix Tables S4, S5)**.

This prioritised set included 6 proteins which were overexpressed in cardiac tissues on mRNA level (**Figure 4**): TIG1 (affecting urinary breakdown products 1-methylxanthine), DNJA4 (affecting urinary breakdown product X-19141), NAR3 (affecting urinary breakdown products n-acetylcitrulline, 4-guanidinobutanoate, and X-23776), SPT20 (affecting urinary breakdown products 1-methylhistamine and X-19141), GLO2 (affecting urinary breakdown products n-methylpipecolate and 3-hydroxyindolin-2-one sulfate), and PLXA1 (affecting urinary breakdown products 1-methylhistamine, glycocholenate sulfate, and 1-methylxanthine) (**Appendix Table S4**). Besides their overexpression in cardiac tissue, these proteins were also overexpressed in at least one other tissue in the body, such as the skeletal muscle and ovaries (**Appendix Table S4**).

**Figure 4.**
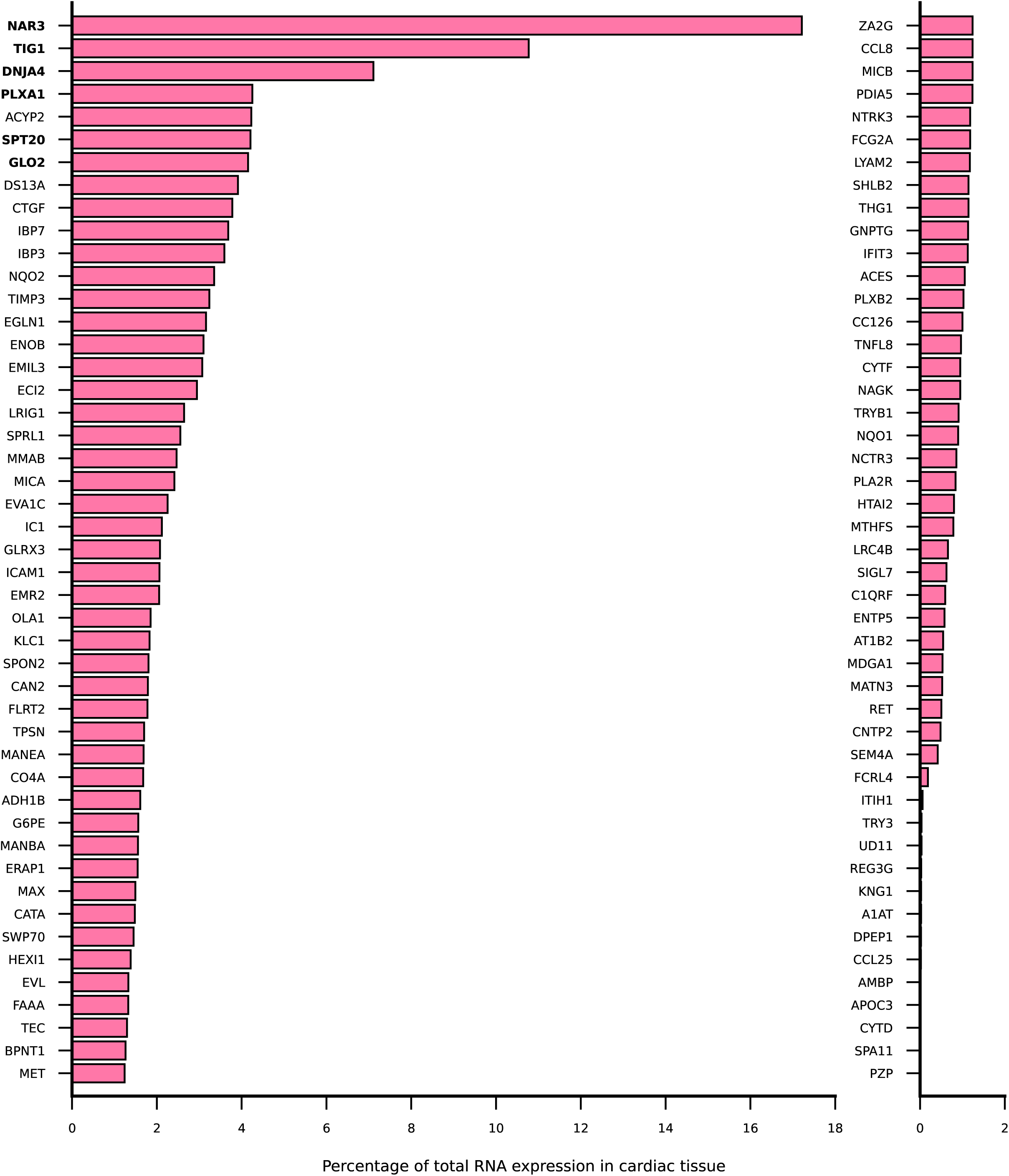
Cardiac mRNA expression of directionally concordant proteins affecting cardiac disease presented as percentage of the total expression across all tissues. N.B. Proteins are identified by their Uniprot label. Proteins displayed in bold font indicate that they are overexpressed in cardiac tissue compared to non-cardiac tissue. Data were sourced from the Human Protein Atlas; please refer to the Methods section for more details.

### Drugged and druggable prioritised proteins

We sourced ChEMBL and BNF to determine druggability of the prioritised proteins. The set of 91 prioritised proteins included 20 drugged proteins, and 12 druggable proteins (step 4 **Figure 1)**. The 20 drugged proteins included seven positive control proteins where the drug indication matched the MR disease association (ACES, EGLN1, ICAM1, NQO2, RET, TRY3, and UD11). For example, we showed that higher levels of ACES increased the risk of both AF and HF, mimicking the protective effect of ACE inhibitor drugs. Furthermore, we identified six proteins targeted by a drug indicated for a cardiac disease distinct from the MR associations (MANBA, NQO1, NTRK3, OLA1, TEC, and TRYB1), identifying compounds relevant for rapid drug repositioning. For example, MANBA is targeted by migalastat used to treat GLA amenable Fabry disease, where the MR analysis supported potential repositioning to treat AF. Additionally, we identified three proteins (AT1B2, DPEP1, and MET) targeted by drug compounds with a registered cardiac side-effect which may be relevant for drug repurposing. Finally, four proteins were targeted by drug compounds without a cardiac indication or side effect, providing further opportunities for drug repurposing. For example, ADH1B (associated with HF) is targeted by nitrefazole which is used to treat alcohol addiction (**Table 1**, **Figure 5, Appendix Tables S7, S8**).

**Figure 5.**
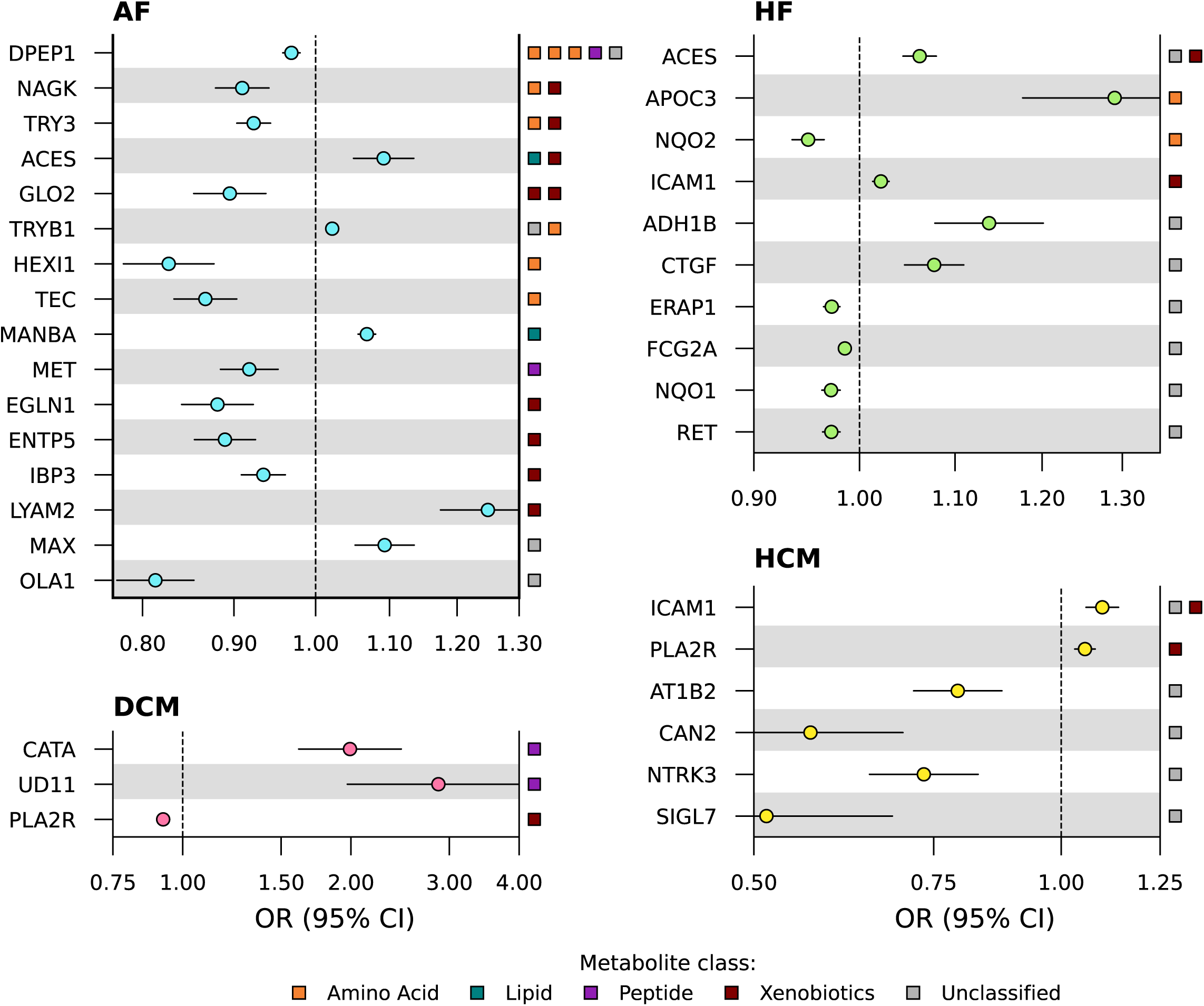
Forest plots of cardiac effects of drugged or druggable prioritised proteins, presented by cardiac disease. N.B. Point estimates are obtained from Mendelian randomisation analyses leveraging genetic data on plasma proteins and cardiac diseases. The metabolite classes of the breakdown products affected by the protein and concordantly associating with the cardiac diseases are indicated on the right y-axis. Genetic associations with the cardiac outcomes were sourced from Nielsen *et al*. (60,620 AF cases)(16), from Shah *et al*. (47,309 HF cases)(17), from Garnier *et al*. (2,719 DCM cases)(18), and from Zhou *et al.* (2,993 HCM cases)(19). For a more detailed description, please refer to the Methods section. Full numerical results can be found in **Appendix Tables S6, S7**. Abbreviations: AF = atrial fibrillation, CI = confidence interval, DCM = dilated cardiomyopathy, HCM = hypertrophic cardiomyopathy, HF = heart failure, OR = odds ratio.

**Table 1.**
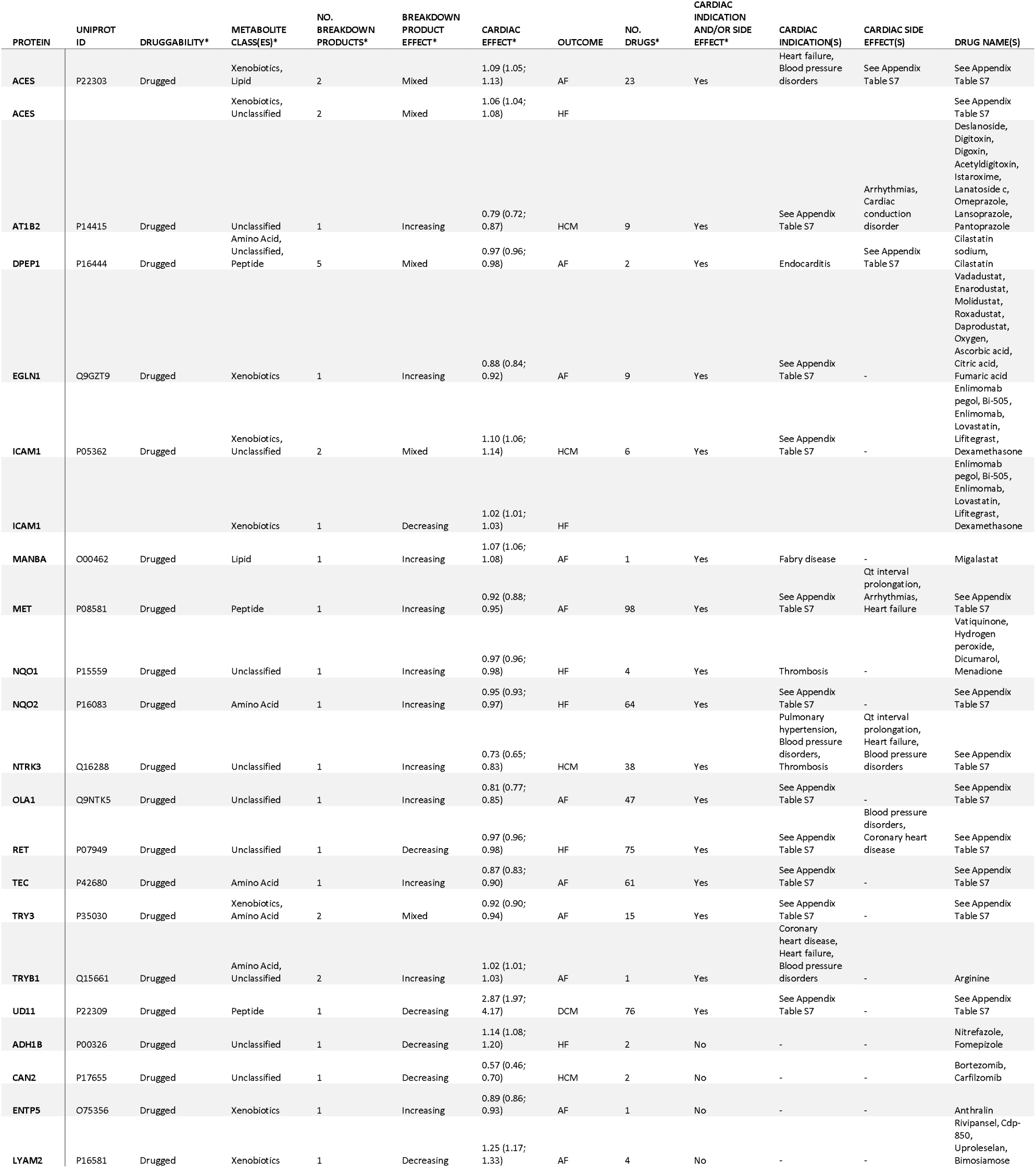
Positive controls and repurposing opportunities: prioritised proteins affected by approved drug compounds. N.B. Characteristics of drugged prioritised proteins and urinary breakdown products associated with one or more cardiac disease(s). Cardiac effect estimates were obtained from *cis* Mendelian randomisation and are presented as odds ratios representing the effect of a one SD increase in protein levels, along with corresponding confidence intervals. The breakdown product effect indicates whether the breakdown product’s effect on cardiac disease is risk-increasing or risk-decreasing, based on *cis* Mendelian randomisation. Druggability was determined based on ChEBML and the British National Formulary. Abbreviations: AF = atrial fibrillation, DCM = dilated cardiomyopathy, HCM = hypertrophic cardiomyopathy, HF = heart failure. More details can be found in Appendix Tables S6, S7.

Irrespective of the known side effects or indications, the identified proteins frequently associated with AF, with many of these proteins (partially) acting through xenobiotic (n=8) and amino acid (n=6) metabolism (**Figure 5**). Specifically, GLO2, ACES, EGLN1 (OR 0.88, 95% CI 0.84; 0.92), LYAM2, IBP3, and ENTP5 affected AF and associated with xenobiotic breakdown product values. Similarly, DPEP1 (OR 0.97, 95% CI 0.96; 0.98), TRYB1, HEXI1, and TEC (OR 0.87, 95% CI 0.83; 0.90) affected AF and associated with amino acid breakdown product values. Higher values of TRY3 (OR 0.92, 95% CI 0.90; 0.94) and NAGK decreased AF risk and associated with both amino acid and xenobiotic breakdown products values.

Most proteins affecting HF associated with unclassified metabolism pathways (n=7). Specifically, increased values of ACES, RET (OR 0.97, 95% CI 0.96; 0.98), ADH1B, ERAP1 (OR 0.97, 95% CI 0.96; 0.98), FCG2A, NQO1, and CTGF affected HF risk and also associated with unclassified urinary breakdown products.

Similarly, drugged and druggable proteins affecting HCM predominantly associated with unclassified metabolism pathways (n=5). Specifically, higher values of ICAM1, NTRK3, CAN2, SIGL7, and AT1B2 affected HCM risk.

For DCM we identified three drugged or druggable proteins with higher values of UD11 and CATA increasing DCM risk and associating with peptide breakdown product prolylglycine, and higher values of PLA2R (OR 0.92, 95% CI 0.91; 0.94) decreasing DCM risk and associating with xenobiotic breakdown product N-methylpipecolate.

### Triangulated proteins with an effect on multiple cardiac traits

We additionally identified eleven pleiotropic proteins associated with multiple cardiac outcomes. Most of these were associated with unclassified and xenobiotic urinary metabolism breakdown products. Specifically, ACES, PLXB2, PLXA1, CYTF, and GNPTG affected AF and HF (**Figure 6**). ICAM1, MICB, and EVA1C affected both HCM and HF, while MATN3 and ACYP2 affected HCM and AF, while PLA2R affected HCM and DCM (**Figure 6, Appendix Table S6**).

**Figure 6.**
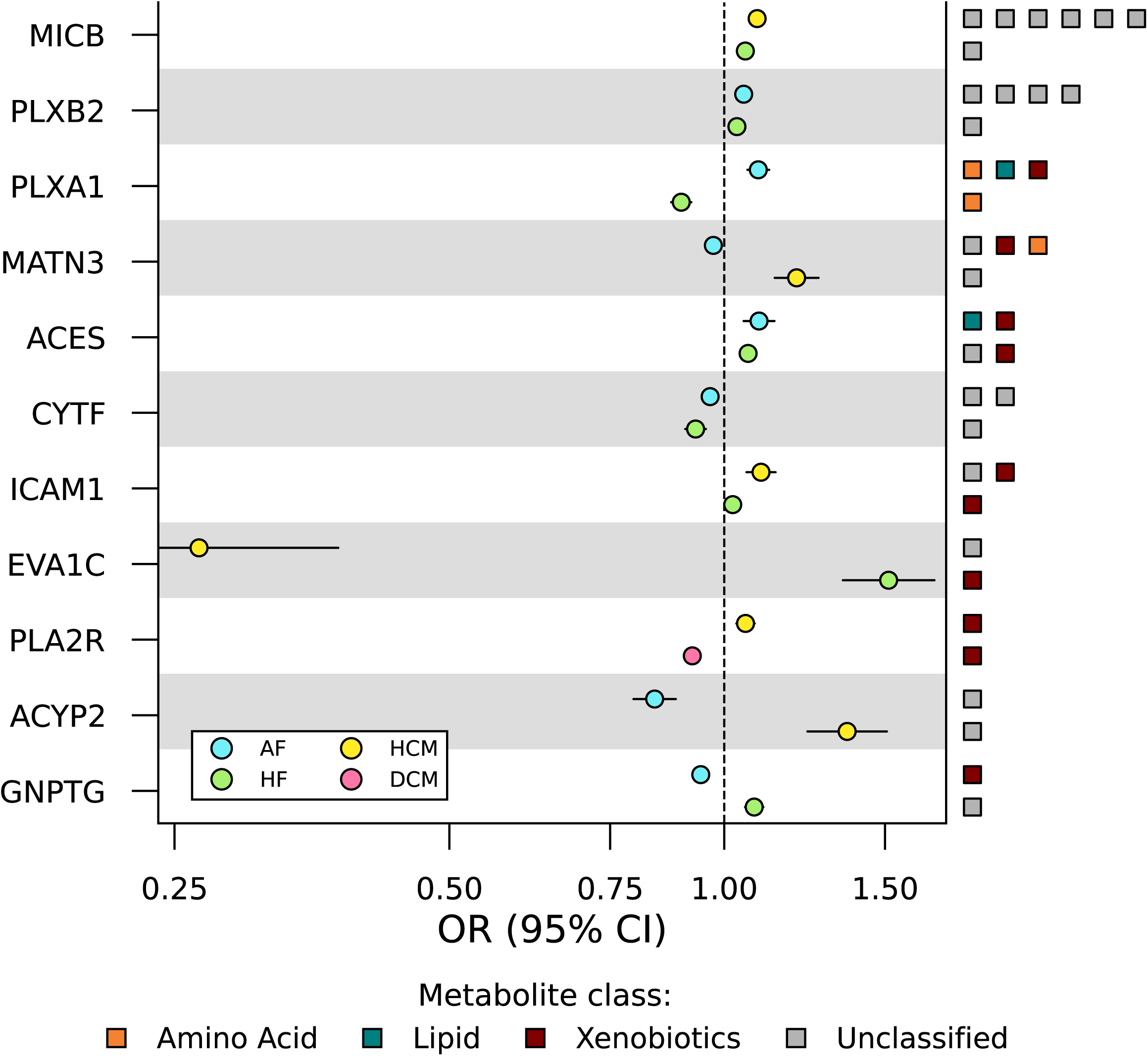
Forest plot of the cardiac effects of prioritised proteins associating with more than one cardiac outcome. N.B. Point estimates are obtained from Mendelian randomisation analyses leveraging genetic data on plasma proteins and cardiac diseases. The metabolite classes of the breakdown products affected by the protein and concordantly associating with each cardiac disease are indicated on the right y-axis. Genetic associations with the cardiac outcomes were sourced from Nielsen *et al*. (60,620 AF cases)(16), from Shah *et al*. (47,309 HF cases)(17), from Garnier *et al*. (2,719 DCM cases)(18), and from Zhou *et al.* (2,993 HCM cases)(19). For a more detailed description, please refer to the Methods section. Full numerical results can be found in **Appendix Table S6**. Abbreviations: AF = atrial fibrillation, CI = confidence interval, DCM = dilated cardiomyopathy, HCM = hypertrophic cardiomyopathy, HF = heart failure, OR = odds ratio.

### Proteins enriched in biological pathways

Based on the Reactome knowledgebase, we found the “antigen presentation: folding, assembly and peptide loading of class I MHC” pathway to be enriched among the triangulated proteins. This pathway involved proteins TPSN (associated with HCM) and ERAP1 (associated with HF) (**Appendix Tables S6, S9**). Enriched pathways in the subset of prioritised proteins per metabolite class and per disease outcome are indicated in **Appendix Figure S4** and **S5**.

### Replication of protein effects on cardiac outcomes

Of the 91 prioritised proteins associating with one or more cardiac traits 56 were available in more than a single study, applying a nominal 0.05 p-value threshold this resulted in a replication of 47 (84%) protein associations with cardiac disease. Applying a multiplicity corrected threshold resulted in 37 (66%) replicated proteins (**Appendix Table S10**). Replication results for protein effects on breakdown product values are described in Supplementary Note 1 (**Appendix Table S11**).

## Discussion

In this study, we used Mendelian randomisation to assess relationships between 954 urinary breakdown products, 1,562 proteins, and four cardiac diseases AF, HF, DCM, and HCM. We found directionally concordant associations between 38 breakdown products, 91 cardiac expressed proteins, and the considered cardiac diseases. The urinary breakdown products are potential biomarkers of altered metabolism pathways informative of the aetiology and consequences of cardiac diseases, and the plasma proteins may be used as drug targets to intervene in these metabolism pathways.

The identified urinary breakdown products associating with cardiac disease predominantly included breakdown products with an origin in amino acids metabolism, xenobiotics metabolism, and of unclassified metabolism. Of the 91 proteins, six were overexpressed in cardiac tissue. We additionally identified eleven pleiotropic proteins with an effect on more than one cardiac outcome. Furthermore, 20 proteins were targeted by an approved drug, which included seven positive control proteins that were targeted by a compound with a cardiac indication matching the MR disease association (ACES, EGLN1, ICAM1, NQO2, RET, TRY3, and UD11), as well as six targets for drugs with a cardiac indication distinct from the MR disease association (MANBA, NQO1, NTRK3, OLA1, TEC, and TRYB1), and three targets for drugs with a known cardiovascular side-effect (AT1B2, DPEP1, and MET) which may be relevant for drug repurposing. For example, we found that the approved compounds nitrefazole and arginine may be beneficially repurposed to treat HF or AF, respectively. Future studies may consider analysis of available (nationwide) prescription data to provide further evidence of efficacy, which may be confirmed by more time consuming and costly randomised controlled trials.

We identified seven amino acids breakdown products associated with AF of which six (cysteinylglycine disulfide, 4-guanidinobutanoate, N-acetylcitrulline, N-acetyl-3-methylhistidine, N-acetylarginine, and N-acetylasparagine) increased the risk of AF, corroborating previous observations of altered amino acid metabolism in AF patients(24). We found 27 proteins associated with these amino acids and AF, including 7 drugged or druggable proteins. For example, we showed that higher values of MANBA (mannosidase beta) increased AF risk, which is supported by recent GWAS associating genetic variants near *MANBA* with AF(25) and by two recent MR studies identifying MANBA as a potential drug target for AF with corresponding effect directions(26, 27). MANBA is targeted by migalastat, a drug used to treat Fabry disease(28), which closely resembles HCM(29), with patients often suffering from cardiac arrhythmias and conduction tissue infiltration, leading to electrical instabilities. An observational study on Fabry disease patients revealed that atrial function was better preserved in those treated with migalastat compared to untreated individuals(30). Through our analyses of urinary breakdown products, we were able to link MANBA to fatty acid metabolism through nonenedioate, with faulty fat metabolism a canonical characteristic of Fabry disease(31).

Jointly, this suggest MANBA may offer therapeutic potential for treatment of AF and related arrythmias through altered lipid metabolism. These insights not only improve our understanding of the metabolic origins of atrial fibrillation but also highlight MANBA as a promising target for intervention. The potential to repurpose migalastat or develop new therapies targeting MANBA offers a novel approach for AF treatment, potentially relevant in patients with metabolic disturbances similar to those observed in Fabry disease. Additionally, we found that higher levels of MET are associated with reduced AF risk. MET is targeted by crizotinib, a cancer drug which has side-effects including arrythmias. Previous research also identified MET as target with repurposing potential for AF treatment(32).

We identified eleven urinary breakdown products associating with HF which included five breakdown products with an origin in amino acids metabolism and five with an unclassified origin. We were able to map these to 27 plasma proteins with a directionally concordant HF effect, which included MICB (MHC class I polypeptide-related sequence B). MICB increased the risk of both HF and HCM, and was linked to seven yet unclassified breakdown products. This closely aligns with our pathway enrichment analysis identifying MHC class I related pathways implicated in HCM (based on protein TPSN) and HF (based on protein ERAP1), which are linked to CD8+ T-cell mediated cardiac remodelling(33), underscoring the potential role of immune modulation in the pathogenesis of these conditions. While these proteins are not yet druggable, our analysis did identify ICAM1 (associating with HCM and HF), RET and NQO2 (both associating with HF), which have already been drugged. Through consideration of urinary breakdown products we were able to associate ICAM1 with 1-methylxanthine and X-18889, where the former is a breakdown product of xanthine, and the oxidation of xanthine has been linked to ICAM1 neutrophil binding capacity(34). Notably, immune cell activation, reflecting systemic but low-grade inflammation, is a likely key pathophysiologic process progressing towards HCM, other cardiomyopathies, and end-stage HF(35). Regorafenib, a RET inhibiting drug that is used for cancer treatment, has registered side effects including myocardial ischemia and hypertension. These side effects support our findings that lower values of RET are associated with a higher risk of HF onset, where the unclassified urinary breakdown product X-23776 was involved in the triangulated association. The GDNF family receptor α–like (GFRAL) acts a receptor for GDF15, which signals through the RET co-receptor. GDF15 was found to be relevant for energy homeostasis and has been identified as a biomarker of cardiovascular and metabolic diseases. There is potential for a new class of drugs based on GDF15-GFRAL-RET, where the signalling pathway is modified(36). Additionally, GDF15 has been recognized as an indicator for HF risk and prediction of hospitalisation for HF or all-cause mortality before(37). The protein NQO2 was associated with a decreased risk of HF, through the amino acid breakdown product indoleacetylglutamine. Prazosin, which acts as an adrenergic receptor alpha-1 antagonist and targets NQO2, has shown potential benefits for HF patients in previous observational studies and trials(38, 39), suggesting prazosin might be a relevant repurposing candidate for HF.

We found five urinary breakdown products and eight proteins associated with DCM risk, including three drugged or druggable proteins, underscoring their potential clinical relevance. For example, we showed that increased values of UD11 increased the risk for DCM and reduced the values of the peptide breakdown product prolylglycine. UD11 is targeted by beta-blockers carvedilol and labetalol (used for HF, hypertension, coronary artery disease and myocardial ischemia), by the diuretics furosemide and bumetanide (used for HF and hypertension) and by acetazolamide, which is not used in practice but has shown benefits in acute decompensated heart failure in a previous clinical trial(40). These findings suggest that modulation of UD11 could be a viable therapeutic strategy in reducing the risk of DCM, providing a rationale for further investigation of UD11 as drug target for treatment for individuals at risk of DCM.

We note that 13 prioritised proteins affected xenobiotic urinary breakdown product values, indicating that these proteins affect xenobiotic metabolism and excretion. For example, we found that ICAM1 associates with the urinary breakdown product value of xenobiotic 1-methylxanthine. This is in line with previous findings indicating that increased expression of ICAM1, which is present on the leukocytes (as well as endothelial cells), facilitates phagocytosis of xenobiotics(41–43).

Despite the robustness of combining multiple MR analyses sourcing various GWAS on both proteins and breakdown products, there are some potential limitations that we wish to discuss. First of all, the genetic data we used was sourced from multiple GWAS which used various high throughput assays, varying in accuracy and analytic scope(44). Instead of directly measuring concentrations, these assays evaluate relative values of proteins or breakdown products. Hence, the magnitude of association is difficult to interpret, and potential follow-up studies are needed to explore compound specific pharmacokinetic and pharmacodynamic properties. Our current analysis predominantly provides information on the putative drug mechanism and whether compounds should be inhibitory or activating. In addition, associations between urinary breakdown products and cardiac disease should not be misinterpreted as implying that urinary breakdown products themselves have a causal effect on disease. Rather we use urinary breakdown products as a proxy for more distal metabolic processes, such as those occurring in or impacting cardiac tissue. Due to relying on aggregate data instead of individual patient data, we could not explore potential influence of variant by environment interactions, for example due to differences in sex or fasting state. However, genetic variants are determined at gametogenesis and therefore MR is protected against most forms of confounding reflecting the presence of a common cause of the genetic variant and the outcome or exposure. Even in the presence of variant by environment interaction, our triangulation approach prioritises results where these interactions are either absent or of minimal importance. This makes our approach conservative, potentially overlooking a number of true associations, but the associations we do identify are robust against environmental influences, which is also indicated by the high degree of replication. Furthermore, GWAS data were mainly obtained from European cohorts, which may limit generalizability to non-European populations. Finally, while we had access to a large sample size GWAS on HF (47,309 cases), we did not have information on HF subtypes, and future studies considering HF subtypes may yield additional insights.

Examining the associations between urinary breakdown products, cardiac disease and proteins in a triangulated way provides a comprehensive and robust analysis. Concordance in results from all three analyses – which are based on distinct sources of data – indicate a more reliable effect. Discordant results however do not necessarily indicate invalid MRs but are potentially indicative of a limited influence of the protein on the metabolism process influencing cardiac disease, or may reflect more intricate biological processes.

In conclusion, 38 metabolism pathways and 91 proteins were identified for involvement with cardiac disease, providing important evidence to support drug development of novel and repurposing targets. Additionally, our manuscript illustrates the value of integrating multi-modal sources of evidence to identify information potentially relevant for disease diagnosis, prognosis, and aetiology.

## Acknowledgements

A preprint version of this manuscript has been deposited at: https://doi.org/10.1101/2024.02.27.24303421. This research has been conducted using the UK Biobank Resource under Application Number 12113. The authors are grateful to UK Biobank participants.

## Author contributions

SD, AFS, and SP designed the study. SD and MV accessed the data, verified the data, and performed analyses. SD drafted the manuscript. All authors provided critical input on the analysis, as well as the drafted manuscript.

## Code availability

Analyses were conducted using Python v3.7.13 (for GNU Linux), Pandas v1.3.5, Numpy v1.21.6, bio-misc v0.1.4, and matplotlib v3.4.3. Code to reproduce figures and tables will be made available upon publication. Genetic summary-level data underpinning each individual MR analysis will be made available at Figshare upon publication.

## Data availability

The individual GWAS data on cardiac outcomes leveraged in this study can be accessed as follows: atrial fibrillation (cases=60,620, total n=1,030,836; https://www.ebi.ac.uk/gwas/publications/30061737), heart failure (cases=47,309, total n=977,323; https://www.ebi.ac.uk/gwas/publications/31919418), dilated cardiomyopathy (cases=2,719, total n=6,980; https://www.ebi.ac.uk/gwas/publications/33677556) and hypertrophic cardiomyopathy (cases=2,993, total n=1,197,200; http://results.globalbiobankmeta.org/pheno/HCM). The GWAS data on urinary breakdown product values can be accessed via https://www.ebi.ac.uk/gwas/publications/31959995 (n=1,627). The individual GWAS data on plasma protein values can be accessed as follows: deCODE (n=35,559, https://www.decode.com/summarydata/), SCALLOP (n=30,931, https://www.ebi.ac.uk/gwas/publications/33067605), Ahola-Olli et al. (n=8,293, https://www.ebi.ac.uk/gwas/publications/27989323), Framingham (n=6,861, https://www.ebi.ac.uk/gwas/publications/21909115), AGES-Reykjavik (n=5,368, https://www.ebi.ac.uk/gwas/publications/35078996), INTERVAL (n=3,301, https://www.ebi.ac.uk/gwas/publications/29875488), Gilly et al. (n=1,328, https://www.ebi.ac.uk/gwas/publications/33303764), and Yang et al. (n=636, https://www.ebi.ac.uk/gwas/publications/34239129).

