## Appendix figures and Supplementary Note for "Multi-omics data integration to identify metabolism pathways and therapeutic targets for cardiac disease"

Content

### **Appendix Figure S1**

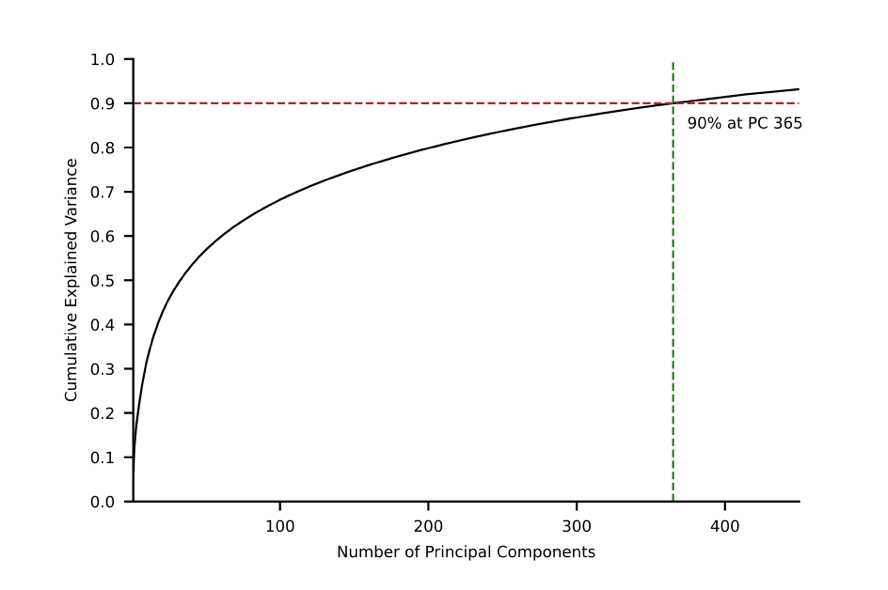

**Figure S1. Cumulative variance plot of principal component analysis**

The number of principal components required to explain 90% of the variance of urinary breakdown product values is depicted. PCA was performed on the Spearman correlation matrix provided by Schlosser *et al*.(15). This study identified and quantified 954 urinary breakdown products among 1,627 participants.

**Appendix Figure S2**

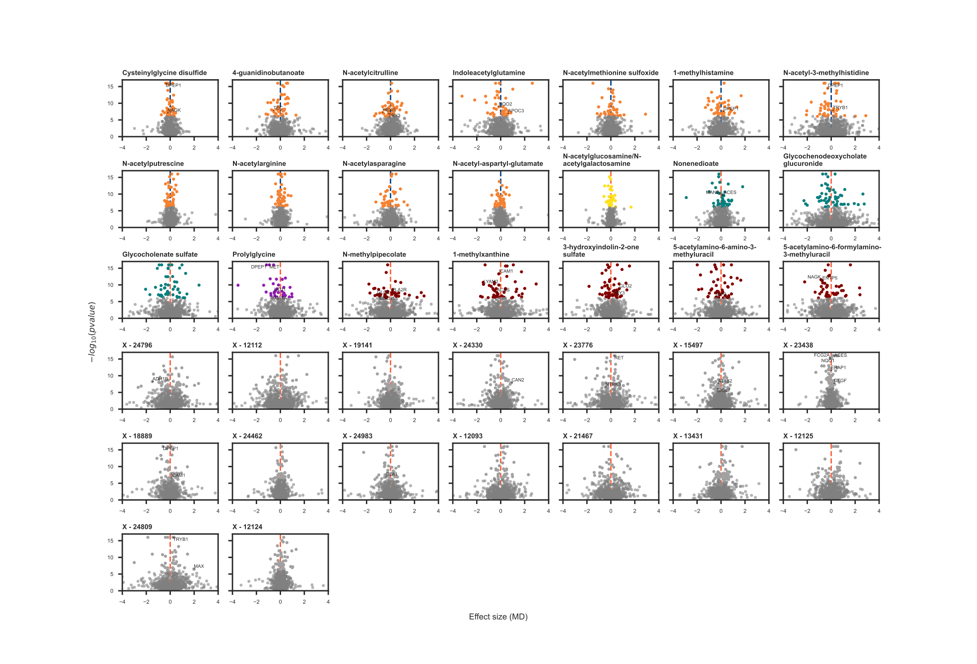

**Figure S2. Volcano plots displaying proteins associated with the breakdown products, presented per breakdown product**

N.B. Presented effect sizes and corresponding p-values are obtained from Mendelian randomisation analyses. Labelled proteins are drugged or druggable, which is defined as proteins targeted by a compound or by a developmental compound (see Methods). The p-values are corresponding to effect estimates obtained from MR estimates and have been truncated to -log10 of 16 for visualisation purposes only. For a more detailed description including descriptions of used data sources and sample sizes, please refer to the Methods section.

Abbreviation: MD = mean difference.

**Appendix Figure S3**

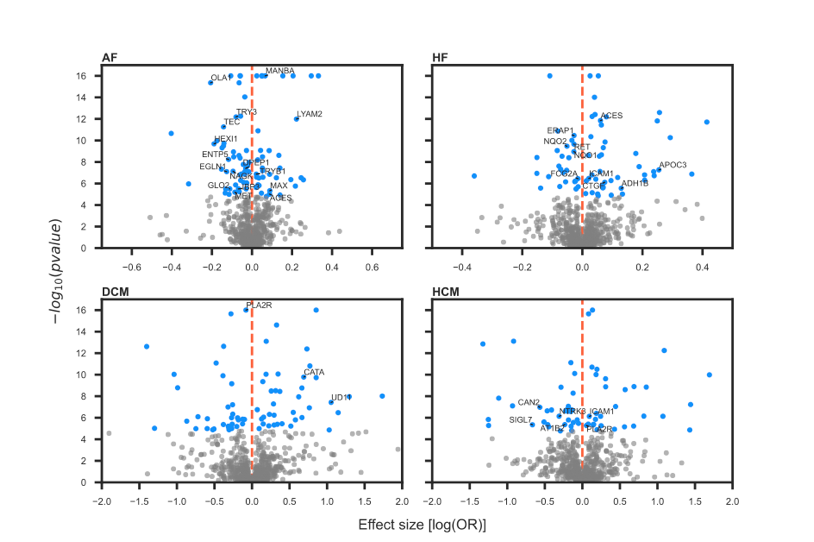

**Figure S3. Volcano plots displaying proteins associated with the cardiac outcomes, presented by cardiac outcome**

N.B. Presented effect sizes and corresponding p-values are obtained from Mendelian randomisation analyses. Labelled proteins are drugged or druggable, which is defined as proteins targeted by a compound or by a developmental compound (see Methods). The p-values are corresponding to effect estimates obtained from MR analyses and have been truncated to -log10 of 16 for visualisation purposes only. For a more detailed description including descriptions of used data sources and sample sizes, please refer to the Methods section.

Abbreviations: AF = atrial fibrillation, DCM = dilated cardiomyopathy, HCM = hypertrophic cardiomyopathy, HF = heart failure, OR = odds ratio.

**Appendix Figure S4**

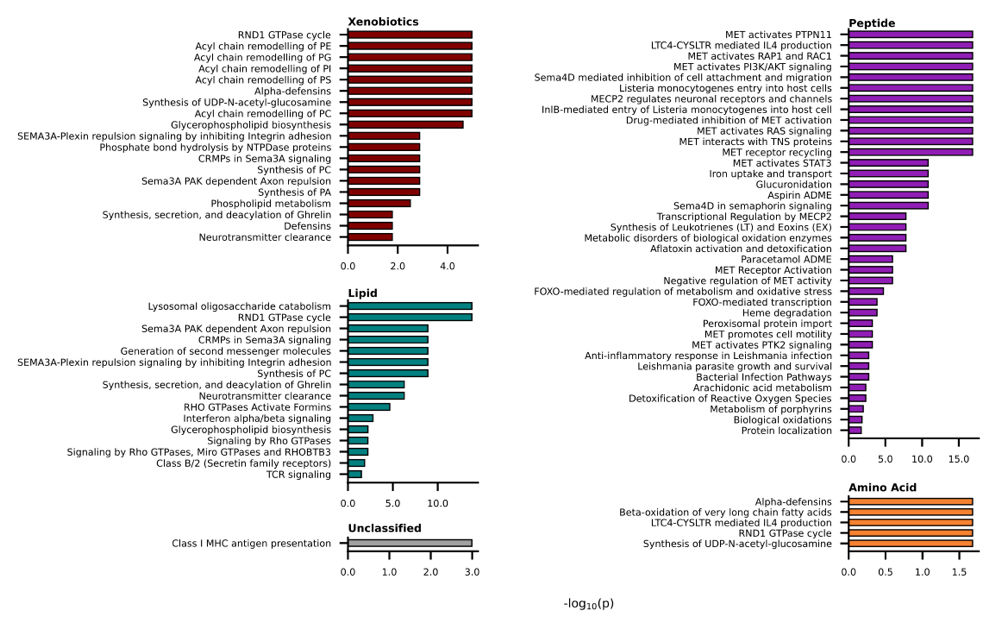

**Figure S4. Enriched Reactome pathways and their corresponding -log10(p-value) per metabolite class**

N.B. Enrichment was based on the Reactome knowledgebase-v85. Prioritised proteins per metabolite class were tested for enrichment against the full set of 1,567 proteins. P-values were adjusted for multiple testing using the Benjamini-Hochberg false discovery procedure.

**Appendix Figure S5**

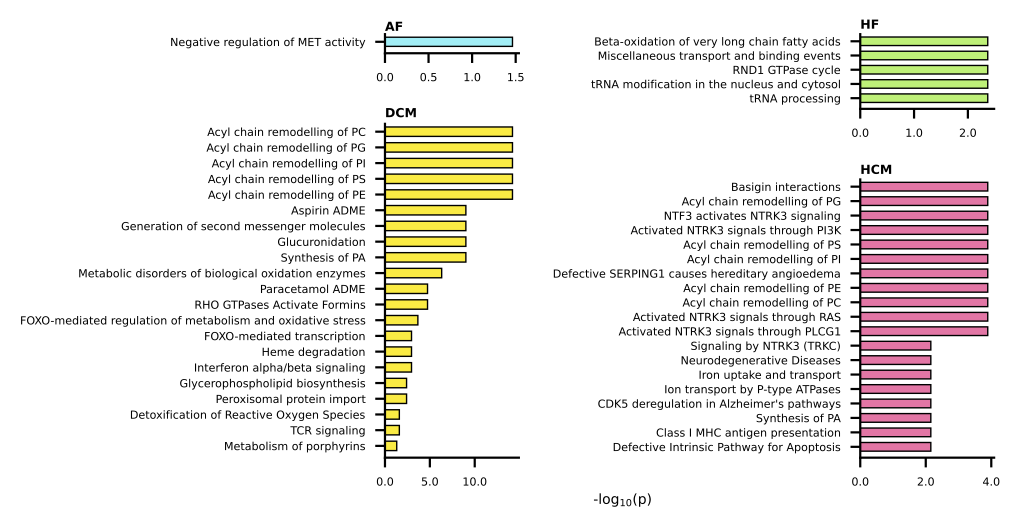

**Figure S5. Enriched Reactome pathways and their corresponding -log10(p-value) per cardiac disease**

N.B. Enrichment was based on the Reactome knowledgebase-v85. Prioritised proteins per cardiac disease were tested for enrichment against the full set of 1,567 proteins. P-values were adjusted for multiple testing using the Benjamini-Hochberg false discovery procedure. Abbreviations: AF = atrial fibrillation, DCM = dilated cardiomyopathy, HCM = hypertrophic cardiomyopathy, HF = heart failure.

### **Supplementary Note 1**

**Methods**

*Cardiac disease GWAS*

Genetic associations with cardiac disease onset were available from Nielsen *et al*. (60,620 AF cases)(16), from Shah *et al*. (47,309 HF cases)(17), from Garnier *et al*. (2,719 DCM cases)(18), and from Zhou *et al.* (2,993 HCM cases)(19). The GWAS on AF by Nielsen *et al*. used six different cohorts, where AF was defined using ICD-9 and ICD-10 codes (427.3 and I48) obtained from local hospitals and outpatient clinics. One cohort defined AF using electronic health records, electrocardiogram results, and/or one in-patient or two-out-patient diagnoses of AF. The percentage of men across cohorts ranged between 49 and 61%. Average age range was not reported by Nielsen *et al*. The GWAS on HF by Shah *et al*. was performed using data from 26 studies. HF was defined using a combination of clinical criteria, diagnostic imaging, physician diagnosis, medication usage, and hospitalization or death records. The ICD codes used are ICD-9 code 428.x and ICD-10 codes I50 and its subcodes. Other specific codes mentioned include ICD-10 codes I11.0, I13.0, I13.2, I25.5, I42.0-9, I50.0-1, I50.9; and ICD-9 codes 4254, 4280-1, 4289. Some studies also used historical ICD-8 codes (427.00, 427.10, 428.99) for HF. The average age of cases across cohorts ranged from 57 to 84 years. The percentage of men within cases across cohorts ranged between 0 and 100%. The GWAS on DCM by Garnier *et al*. was performed on five populations of European ancestry (France, Germany, USA, Italy, and UK) where sporadic DCM was diagnosed according to standard criteria by reduced ejection fraction and enlarged left ventricular end-diastolic volume/diameter in the absence of any obvious pathology. The average age at inclusion across populations ranged from 43 to 55 years. The percentage of men within cases across populations ranged from 66.6 to 82.1%.

The GWAS on HCM by Zhou *et al*. was obtained from a collaborative network of 23 biobanks where HCM was defined using ICD-9 and ICD-10 codes (425.1 and I42.1), or through other available health data, such as self-report data. The average age across cohorts ranged from 40 to 70 years. The range of sex representation was not reported by Zhou *et al*.

*Proteomic GWAS*

Genetic associations with plasma protein values were available from the following eight GWAS: deCODE (SomaLogic assay, n=35,559)(47), SCALLOP (Olink assay, n=30,931)(48), Ahola-Olli et al. (BioRad assay, n=8,293)(49), Framingham (Luminex assay, n=6,861)(50), AGES-Reykjavik (SomaLogic assay, n=5,368)(51), INTERVAL (SomaLogic assay, n=3,301)(52), Gilly et al. (Olink assay, n=1,328)(53) and Yang et al., (SomaLogic assay, n=636)(54).

*Mendelian randomisation assumptions and model selection framework*

MR analysis relies on three core assumptions. First, genetic variants are assumed to be strongly associated with the exposure variable (relevance). Second, genetic variants are assumed to have no common cause with either the exposure or outcome variables (independence). Third, the effect of genetic variants on the outcome is assumed to be through the exposure variable only (exclusion restriction).

A model selection framework was used to identify the MR model most supported by the data. In the absence of meaningful heterogeneity, the model selection framework favours the IVW method over MR-Egger estimator because it generally yields more accurate estimates (i.e. higher precision) but selects MR-Egger regression in case it provides a demonstratively better fit to the data(55). Within the framework, the goodness of fit to the data is expressed as a Q-statistic.

*mRNA expression and enrichment*

Cardiac mRNA expression was sourced from the Human Protein Atlas (HPA), utilizing the consensus expression derived from normalised transcripts per million (nTPM) values from three independent transcriptomics datasets. These datasets include GTEx (Genotype-Tissue Expression), which provides a public resource of human gene expression and regulation across multiple tissue types; Fantom5 (Functional Annotation of the Mammalian Genome 5), known for its detailed atlas of human tissues integrating promoters and enhancers expression profiles; and HPA's own dataset, which provides data focused on protein expression across various human tissues, including a variety of cardiac tissues such as myocardium, atrial appendages, and ventricles. Overexpressed genes were identified by comparing cardiac expression with average expression in other tissues, testing against a standard normal quantile of 1.96.

*Druggability*

Druggability of proteins was obtained from ChEMBL and the British National Formulary (BNF). The BNF is a collaborative effort of the British Medical Association and the Royal Pharmaceutical Society that sources information from drug inserts, literature, regulatory agencies, and professional organizations. ChEMBL is a source of information related to clinically used drug targets (from US FDA-approved drugs) and potential targets that are currently under investigation, as described by Finan and colleagues(56).

**Results**

*Replicating protein associations with breakdown products*

Of the 91 prioritised proteins 54 were available in more than a single study allowing for MR analyses of protein on urine breakdown product effects, representing 59% (95% CI 49; 70) of all the triangulated protein associations. Breakdown product associations were replicated for 50 proteins, when applying a nominal p-value of 0.05. Using a more stringent p-value of 9.26×10^-4^ (0.05 divided by the number of proteins that were available in more than one study) allowed for 41 replications of protein effects on breakdown products.

**Appendix Tables S7 and S8**

In this paper, for the construction of appendix tables S7 and S8, the following oncologic indications and side effects were grouped under the general category of 'Cancers':

Adenoma, islet cell, Ameloblastoma, Astrocytoma, Blast crisis, Carcinoma, Castleman disease, Chordoma, Craniopharyngioma, Diffuse intrinsic pontine glioma, Ependymoma, Esthesioneuroblastoma, olfactory, Gastrinoma, Gestational trophoblastic disease, Ganglioneuroblastoma, Glioblastoma, Glioma, Glaucoma, Granuloma, lethal midline, Hemangiopericytoma, Hemangioendothelioma, Hemangioendothelioma, epithelioid, Hematoma, Hepatoblastoma, Hereditary breast and ovarian cancer syndrome, Hodgkin disease, Insulinoma, Leukemia, Lymphoma, Malignant carcinoid syndrome, Mastocytosis, Medulloblastoma, Melanoma, Meningioma, Mesothelioma, Metastatic colorectal cancer, Multiple myeloma, Mycosis fungoides, Myelodysplastic syndromes, Myeloproliferative disorder, Neoplasms, Neurilemmoma, Neuroblastoma, Neurofibroma, plexiform, Oligodendroglioma, Optic nerve glioma, Papilloma, inverted, Pinealoma, Plasmacytoma, Primary myelofibrosis, Pseudomyxoma peritonei, Retinoblastoma, Ros1-positive advanced non-small cell lung cancer, Sarcoma, Sezary syndrome, Seminoma, Smoldering multiple myeloma, Somatostatinoma, Teratoma, Thymoma, Tumors, and Waldenstrom macroglobulinemia.
